# Global Disparities in Neuroimaging Research for Mental Health Conditions

**DOI:** 10.64898/2026.01.27.26344057

**Authors:** Tatum Connell, Caio B. Casella, Nathalia B. Esper, Nim Tottenham, Mark Tomlinson, Agustin Ibanez, Antonis A. Kousoulis, Soraya Seedat, Jason Bantjes, Brandon A. Kohrt, Matias Irarrazaval, Stephanie Ameis, Luis Augusto Rohde, Dan J. Stein, Paul Thompson, Pedro Mario Pan, Zul Merali, Pedro Antonio Valdés-Sosa, Christian Kieling, Michael P. Milham, Zeina Mneimneh, Giovanni A. Salum

## Abstract

**Background:** Scientific research remains disproportionately grounded in data from high-income countries (HICs), yet efforts to map the distribution of neuroimaging findings by income levels remain limited.

**Methods:** Using data from the ENIGMA Consortium, we conducted a systematic quantitative synthesis of 83 publications across nine psychiatric and neurological conditions, analyzing T1-weighted structural MRI data from 16,086 cases in 27 countries. Representation was mapped using World Bank income classifications and World Health Organization (WHO) regions.

**Results:** HICs contributed 90.5% of cases; upper-middle-income countries 7.9%; lower-middle-income countries 1.6%; and low-income countries none. Geographically, 85% of cases originated from North America and Europe, while Africa, South-East Asia, and the Eastern Mediterranean were underrepresented. Supplemental analyses of other datasets (Brain Growth Charts; fMRI meta-analysis) revealed similar disparities.

**Conclusions:** Equitable neuroimaging science is critical to inform practice and policy decision-making that is context specific. This requires targeted investment in infrastructure, data sharing, and participation from underrepresented regions.

## Introduction

There is a broad pattern in scientific research that relies on Western, educated, industrialized, rich, and democratic (WEIRD) populations as the default study sample^1^. This reflects the longstanding 10/90 gap, where only an estimated 10% of global health research funding addresses the health needs of the poorest 90% of the world’s population^2^. Biases in data representation could severely hinder scientific progress and limit the generalizability of findings since sociocultural, genetic, and environmental factors substantially influence human development^3^. Computational neuroimaging models are highly dependent on how data are represented and require broad diversity to build accurate and robust precision models^4^. For neuroscience to generate real-world impact, it must actively incorporate real-world diversity, accounting for population heterogeneity, geographic distribution, and context^5,6^. While some fields have advanced characterization of the extent to which our evidence relies on biased estimates^7,8^, this is still not well quantified in the neuroimaging field, particularly when it pertains to understanding mental health conditions.

Previous evidence on the topic of biased estimates in neuroimaging research is limited. A study investigating adolescent pediatric depression quantified that 82% of studies on teenage depression originate from high-income countries^9^. Major neuroimaging datasets, such as the ABCD Study, Human Connectome Project, and UK Biobank, are overwhelmingly composed of white participants, with up to 95% of data in some cohorts drawn from this demographic^3^. Similarly, a recent large-scale effort to develop lifespan brain charts that compiled over 120,000 MRI scans acknowledged that the dataset was disproportionately drawn from individuals of European and North American ancestry, limiting its global applicability^10^.

One major effort to address global representation in neuroimaging is the Enhancing Neuroimaging Genetics through Meta-Analysis (ENIGMA) Consortium. Launched in 2009, it is the largest global collaborative platform in imaging and genetics^11^. It brings together over 2,000 scientists across 47 countries to investigate brain structure, function, and risk for psychiatric, developmental, and neurological conditions. ENIGMA pools existing datasets from around the world and coordinates harmonized analyses across more than 50 active working groups, each focused on a specific psychiatric, neurological, or developmental condition, including cross-disorder analyses^12^. These standardized protocols allow researchers to combine large, diverse datasets while maintaining consistent data quality across sites. Given its broad scope and commitment to diversity, ENIGMA constitutes one of the few initiatives that allows for consistent extraction and comparison of geographic and economic representation, making it an ideal source for quantifying data representation patterns in neuroimaging.

To our knowledge, no study has systematically examined the availability of structural MRI data across multiple mental health conditions globally. Existing analyses often center on specific populations or disorders, overlooking broader structural patterns of inclusion and exclusion within the field^3,7,13^. To address these critical gaps, the present study investigates disparities in data availability for structural MRI data for mental health and neurological conditions between HICs and LMICs. Building on data from the ENIGMA Consortium^10^, here we conduct a systematic quantitative synthesis to quantify and map structural MRI data availability according to the World Bank Income Classification^14^ and the World Health Organization (WHO)^15^ regions. We supplemented our analysis by examining whether the geographic and economic disparities identified in ENIGMA data were consistent across other large-scale neuroimaging initiatives. Specifically, we analyzed data from the Brain Charts Consortium^10^, which assessed structural imaging markers of typical development, thereby avoiding bias toward mental health populations, using the same MRI modality. We also included a meta-analysis by Tamon et al.^16^, which combined 243 studies of different modality (task-based fMRI studies) in ADHD and ASD, helping to mitigate potential bias related to willingness to participate in a research consortium.

## Methods

We conducted a systematic quantitative synthesis of ENIGMA Consortium publications that used structural T1-weighted MRI in clinical populations. To identify eligible studies, we searched PubMed using the terms (ENIGMA) AND (“Structural MRI” OR “T1”), yielding 83 results. Additionally, we manually reviewed working group pages on the ENIGMA website to capture relevant studies not identified by the database search.

Studies were included if they: (1) involved structural T1-weighted MRI data; (2) focused on a clinical population diagnosed with one of nine psychiatric or neurological conditions: Attention-Deficit/Hyperactivity Disorder (ADHD)^17^, Anorexia Nervosa^18^, Autism Spectrum Disorder (ASD)^19^, Epilepsy^20^, Generalized Anxiety Disorder (GAD)^21^, Obsessive-Compulsive Disorder (OCD)^22^, Social Anxiety Disorder (SAD)^23^, Schizophrenia^24^, and Substance Use Disorder^25^; (3) were conducted under the ENIGMA Consortium; and (4) reported study-level information, including participant demographics and sample information. When multiple studies examined the same condition or sample, we selected the one with the larger sample size. We also excluded conditions (Major Depressive Disorder^26^, Conduct Disorder^27^, and Bipolar Disorder^28^) without site-specific data or those from multi-country cohorts where participant numbers could not be attributed to individual countries. We consolidated all substance use disorders (cannabis, cocaine, alcohol, methamphetamine, and nicotine use disorders) into a single “Substance Use Disorder” category.

We reviewed the full text and supplemental materials for each included publication to extract study-level data. Information was entered into a structured Excel database and included: contributing sites, ENIGMA working group, clinical condition, age group, and total sample size. We recorded the number of participants, mean age with standard deviation, and percentage of female participants for both cases and controls. We also extracted country-level data for each contributing site, including country name, World Bank income classification, and WHO region.

All analyses were conducted on clinical (case) participants using R (version 4.4.3) and the ggplot2^29^, sf^30^, and rnaturalearth^31^ packages. First, we standardized the World Bank income classifications (2023) into abbreviated categories: high-income countries (HIC), upper-middle-income countries (UMIC), lower-middle-income countries (LMIC), and low-income countries (LIC). We conducted additional analyses to characterize temporal trends in global representation by examining publication years and regional participation over time. We also systematically documented studies that could not be included due to insufficient geographic detail to estimate the scope of potentially excluded data.

We focused specifically on T1-weighted structural MRI data because: (1) it represents the most commonly acquired and standardized neuroimaging sequence across global sites, (2) it allows for consistent morphometric analyses across diverse scanner types and field strengths commonly found in different resource settings, and (3) it provides the foundation for most structural brain analyses in psychiatric research, making it the optimal choice for assessing global representation patterns.

We aggregated the total number of cases for each clinical condition and calculated the proportion contributed by each income group and WHO region. Countries were categorized into one of four income levels and assigned to one of the six WHO regions: African Region (AFR), Region of the Americas (AMR), Eastern Mediterranean Region (EMR), European Region (EUR), South-East Asia Region (SEAR), and Western Pacific Region (WPR). We calculated the percentage of total cases represented by each income level and WHO region. Results were compiled into a summary table (Table 1). Heatmaps were generated to visualize the geographic distribution of ENIGMA data at the country level. Figure 1 presents the number of contributing study sites per country, aggregated across all nine conditions. Supplemental maps provide additional detail: Figure S1 shows structural MRI case counts by country for each condition; Figure S2 displays study site counts by country for each condition; and Figure S3 presents total case counts by country across all conditions, grouped into distinct categories (0, 1–50, 51–100, 101–200, 201–500, 501-1000, >1000).

**Figure 1.**
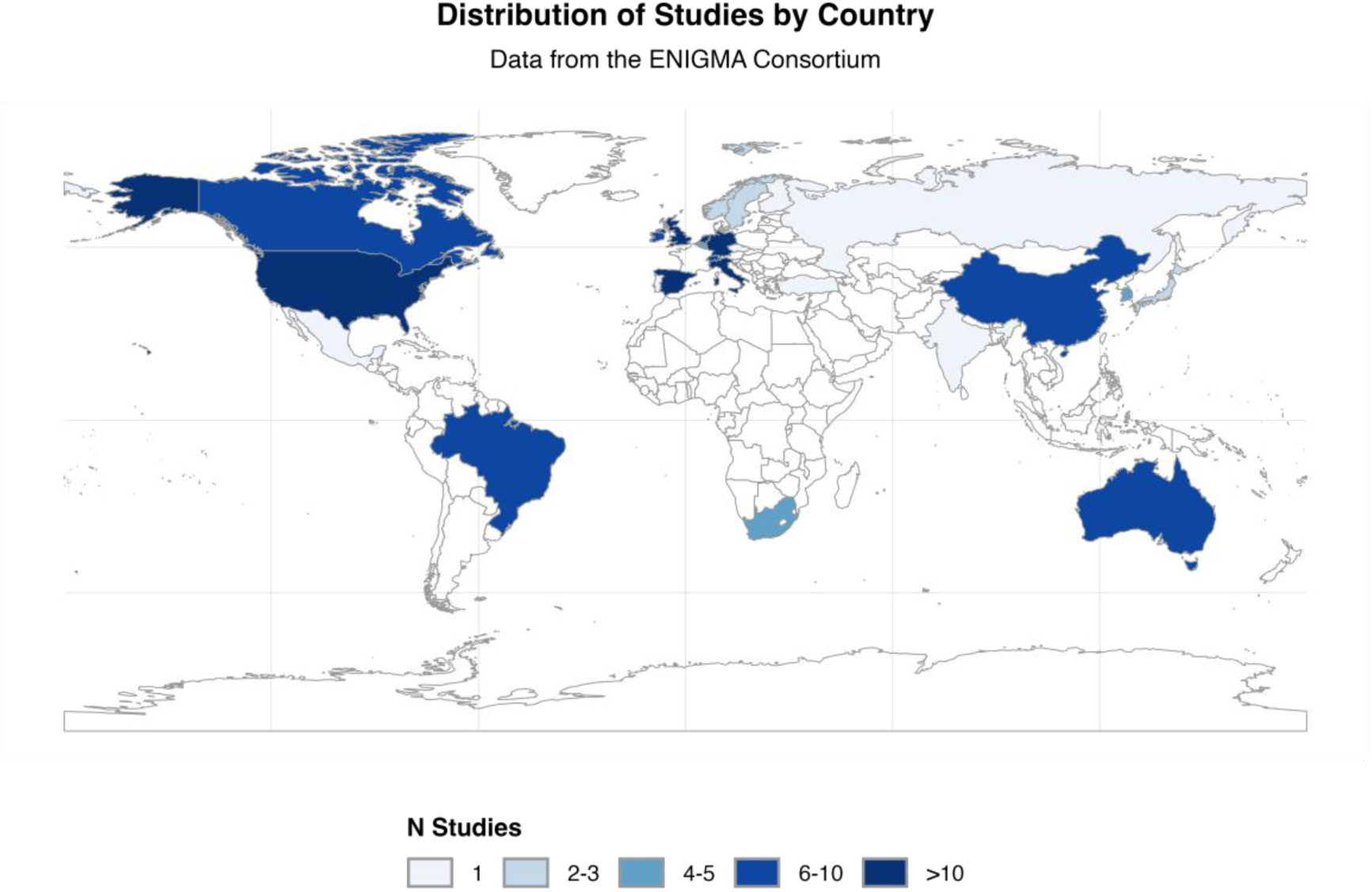
Distribution of Studies by Country. The figure shows the geographic distribution of studies included from the ENIGMA Consortium database across combined conditions: ADHD, Anorexia, Autism, Epilepsy, Generalized Anxiety Disorder, Obsessive Compulsive Disorder, Social Anxiety Disorder, Schizophrenia, and Substance Use Disorder. Color intensity represents the number of studies per country.

**Table 1.** Distribution of Cases by World Bank Income Level and WHO Region.

|  | World Bank Income Level |  |  |  | WHO Region |  |  |  |  |  |  |
| --- | --- | --- | --- | --- | --- | --- | --- | --- | --- | --- | --- |
| Condition | HIC | UMIC | LMIC | LIC | AFR | AMR | EMR | EUR | SEAR | WPR | Total N |
| **ADHD** | 1806<br>(93.4%) | 127<br>(6.6%) | .. | .. | .. | 820<br>(42.4%) | .. | 963<br>(49.8%) | .. | 150<br>(7.8%) | **1933** |
| **Anorexia** | 685<br>(100.0%) | .. | .. | .. | .. | 158<br>(23.1%) | .. | 527<br>(76.9%) | .. | .. | **685** |
| **Autism** | 1818<br>(99.2%) | 15<br>(0.8%) | .. | .. | .. | 1131<br>(61.7%) | .. | 702<br>(38.3%) | .. | .. | **1833** |
| **Epilepsy** | 1764<br>(82.1%) | 385<br>(17.9%) | .. | .. | .. | 657<br>(30.6%) | .. | 1209<br>(56.3%) | .. | 283<br>(13.2%) | **2149** |
| **GAD** | 1005<br>(98.5%) | 15<br>(1.5%) | .. | .. | .. | 858<br>(84.1%) | .. | 162<br>(15.9%) | .. | .. | **1020** |
| **OCD** | 1779<br>(78.1%) | 234<br>(10.3%) | 265<br>(11.6%) | .. | 23<br>(1.0%) | 568<br>(24.9%) | .. | 849<br>(37.3%) | 441<br>(19.4%) | 397<br>(17.4%) | **2278** |
| **SAD** | 1017<br>(91.2%) | 98<br>(8.8%) | .. | .. | 22<br>(2.0%) | 469<br>(42.1%) | .. | 605<br>(54.3%) | .. | 19<br>(1.7%) | **1115** |
| **Schizophrenia** | 2479<br>(88.4%) | 324<br>(11.6%) | .. | .. | 90<br>(3.2%) | 742<br>(26.5%) | .. | 1099<br>(39.2%) | 126<br>(4.5%) | 746<br>(26.6%) | **2803** |
| **Substance** | 2198<br>(96.8%) | 72<br>(3.2%) | .. | .. | 72<br>(3.2%) | 1785<br>(78.6%) | .. | 316<br>(13.9%) | .. | 97<br>(4.3%) | **2270** |
| **Total ENIGMA** | 14551<br>(90.5%) | 1270<br>(7.9%) | 265<br>(1.6%) | .. | 207<br>(1.3%) | 7188<br>(44.7%) | .. | 6432<br>(40%) | 567<br>(3.5%) | 1692<br>(10.5%) | **16086** |
| **Supplemental Analysis** |  |  |  |  |  |  |  |  |  |  |  |
| **Bethlehem et al. (2022)** | 118223<br>(95.4%) | 4367<br>(3.5%) | 1394<br>(1.1%) | .. | 137<br>(0.1%) | 55598<br>(44.8%) | .. | 59985<br>(48.4%) | 1394<br>(1.1%) | 6870<br>(5.5%) | **123984** |
| **Tamon et al. (2024)** | 5689<br>(98.3%) | 96<br>(1.7%) | .. | .. | .. | 2119<br>(36.6%) | .. | 3078<br>(53.2%) | .. | 588<br>(10.2%) | **5785** |

Data shown as N (%). HIC=high-income countries. UMIC=upper-middle-income countries. LMIC=lower-middle-income countries. LIC=low-income countries. AFR=African Region. AMR=Region of the Americas. EMR=Eastern Mediterranean Region. EUR=European Region. SEAR=South-East Asia Region. WPR=Western Pacific Region. ..=no data available. ADHD: attention-deficit/hyperactivity disorder; GAD: Generalized Anxiety Disorder; OCD: Obsessive Compulsive Disorder; SAD: Social Anxiety Disorder

To visualize disparities between the ENIGMA sample and the global population, we created bar plots comparing the proportion of structural MRI cases with 2023 population estimates across World Bank income groups and WHO regions (Figure 2). We normalized the population counts to percentages and plotted alongside the share of ENIGMA cases from each category to highlight representation gaps.

**Figure 2.**
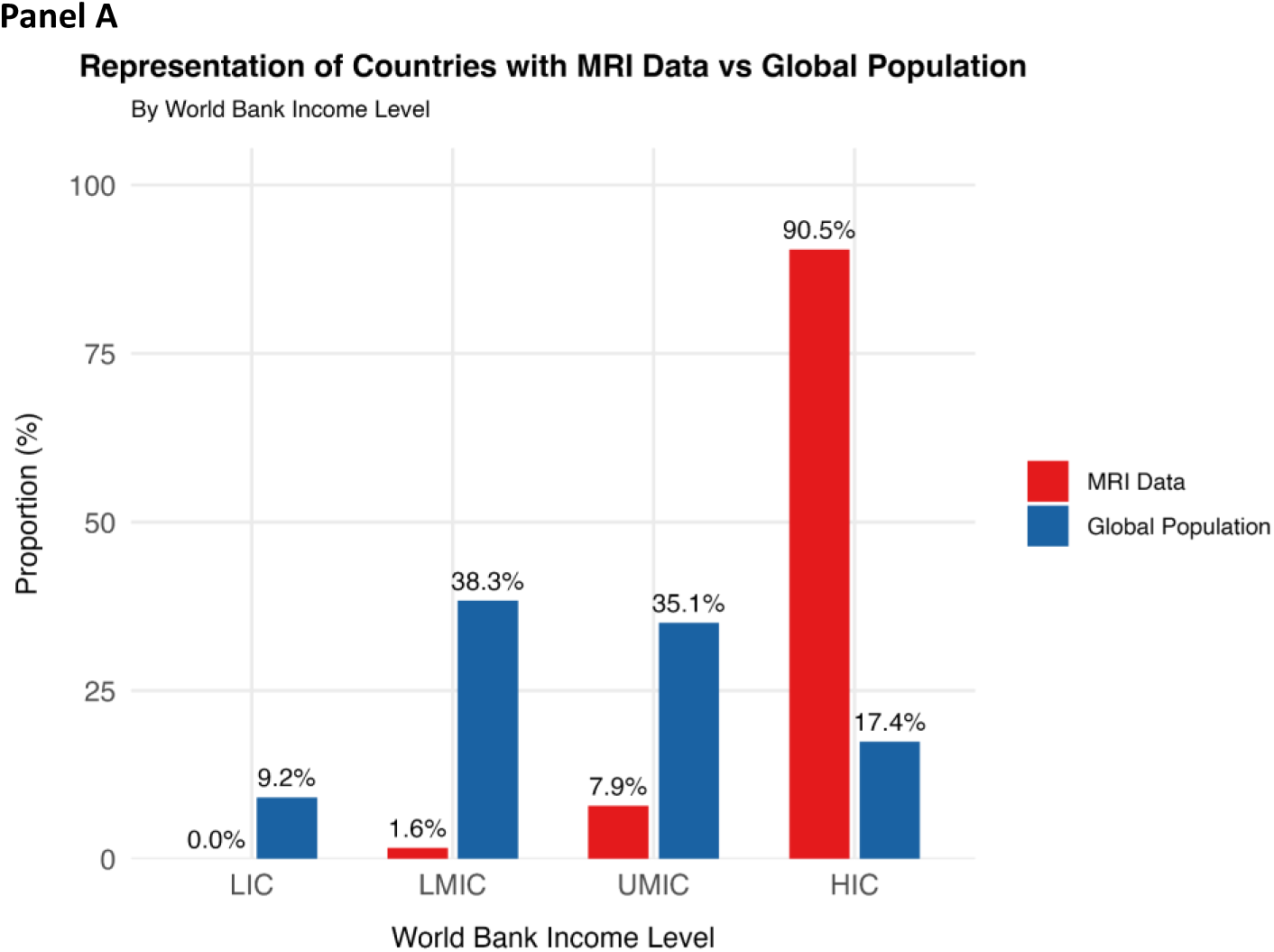
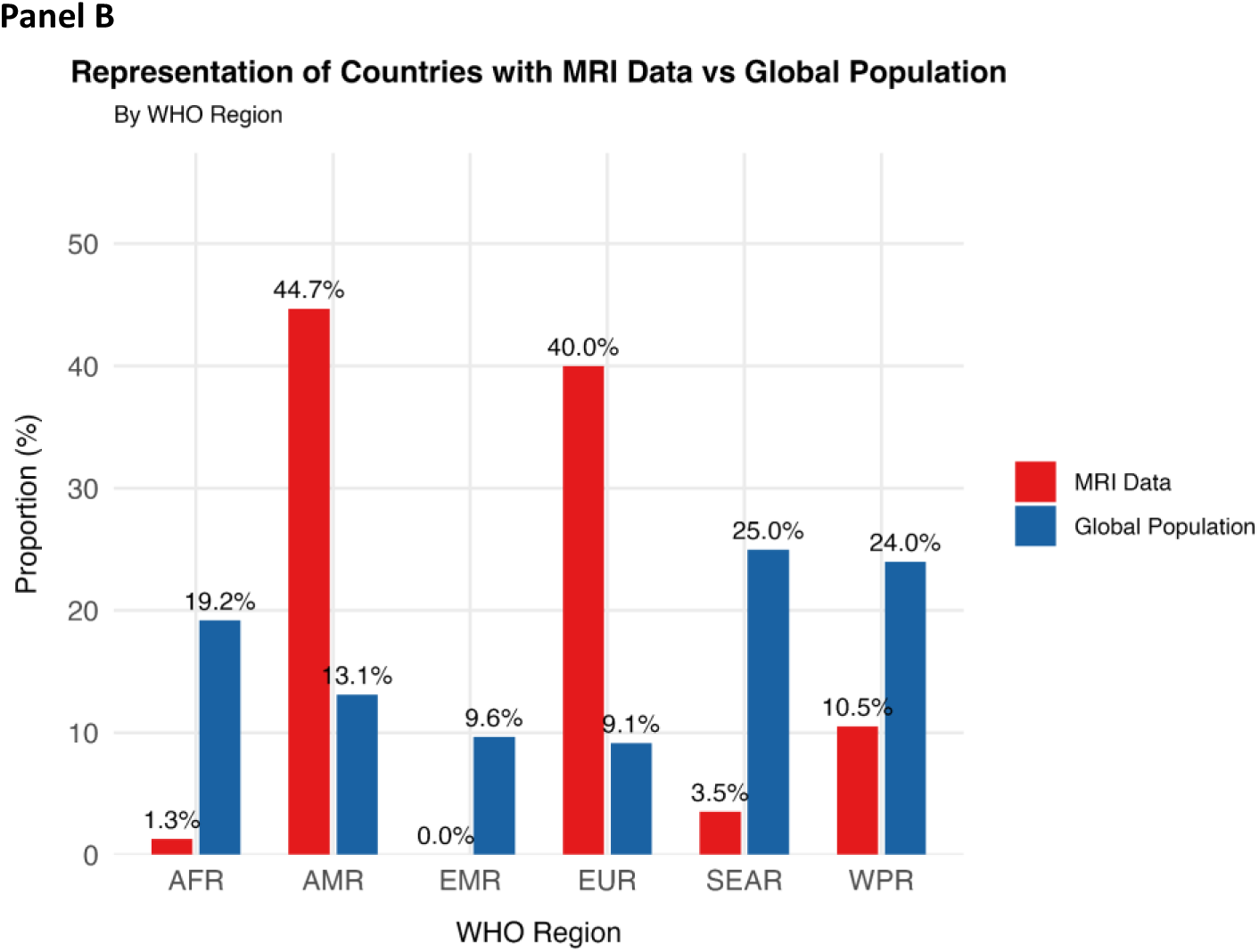
Representation of Countries with MRI Data vs Global Population (World Bank Income Levels). This figure shows the proportion of countries with structural MRI data available using ENIGMA data compared to the global population, grouped by World Bank income level (high, upper-middle, lower-middle, and low income). HIC=high-income countries. UMIC=upper-middle-income countries. LMIC=lower-middle-income countries. LIC=low-income countries. This figure shows the proportion of countries with structural MRI data available using ENIGMA data compared to the global population, grouped by the World Health Organization (WHO) regions. AFR=African Region. AMR=Region of the Americas. EMR=Eastern Mediterranean Region. EUR=European Region. SEAR=South-East Asia Region. WPR=Western Pacific

Finally, we included a supplemental analysis to contextualize our ENIGMA findings within the broader neuroimaging field. We used data from the Brain Charts Consortium^10^, which aggregated over 120,000 MRI scans to develop lifespan brain charts, and Tamon et al. (2024)^16^, a meta-analysis of task-based fMRI studies on ADHD and ASD. For each study, we identified contributing countries from the published datasets and mapped them to World Bank income classifications and WHO regions using the same methodology applied to ENIGMA data. For the Tamon et al (2024) dataset, we included only case participants in the totals and analyses.

## Results

Our systematic quantitative synthesis aggregated 16,086 structural MRI cases across nine mental health and neurological conditions from the ENIGMA Consortium. The distribution of cases demonstrated pronounced disparities across World Bank income levels and WHO regions. These variations are further illustrated in heatmaps of case distribution (Figure S1), with dense case concentrations in North America and Western Europe.

HICs contributed 90.5% (N=14,551) of all structural MRI cases, while UMICs contributed 7.9% (N=1,270) and LMICs contributed only 1.6% (N=265). No data from LICs were identified for any condition studied. Across the dataset, 27 unique countries contributed data, and the number of contributing sites per condition ranged from 22 to 57. As shown in Figure 1, study sites were overwhelmingly concentrated in HICs, with over 100 based in the United States. OCD, epilepsy, and schizophrenia had the widest geographic reach, with studies from 14 to 16 countries, while GAD and substance use disorder were the most limited.

Disparities in income representation varied substantially across conditions. OCD was the only condition with increased LMIC participation, with 11.6% (N=265) of OCD cases originating from these countries. All other conditions were represented exclusively by data from HICs and UMICs, despite their well-established global prevalence. Epilepsy demonstrated the most balanced income distribution, with 17.9% (N=385) of epilepsy cases contributed by UMICs. Anorexia nervosa was the most economically restricted condition, with 100% of data derived from HICs.

The distribution across WHO regions revealed that most data originated from the Region of the Americas (AMR) (N=7,188, 44.7%) and the European Region (EUR) (N=6,432, 40.0%); together, these two regions accounted for 84.7% of the total dataset. The Western Pacific Region (WPR) contributed 1,692 cases (10.5%), followed by the South-East Asia Region (SEAR) with 567 cases (3.5%), and the African Region (AFR) with 207 cases (1.3%). No data was available from the Eastern Mediterranean Region (EMR). Figure 2 (Panel A) shows that low-and lower-middle-income countries make up nearly 48% of the world population yet contributed less than 2% of ENIGMA cases. Similarly, Panel B highlights the mismatch between the global population and study representation across WHO regions: Europe and the Americas contribute over 80% of ENIGMA cases despite comprising less than 23% of the world population, while Africa and South-East Asia represent nearly 45% of the global population but account for under 5% of cases.

Geographic diversity varied by condition. No condition was represented across all six WHO regions. OCD showed the widest spread, with data from 18 countries spanning five WHO regions. Schizophrenia also had data from five regions. Substance use disorder and SAD were represented in four regions. In contrast, anorexia nervosa, ASD, and GAD were the most geographically restricted, with data mainly from HICs in just two regions. These patterns underscore the critical underrepresentation of Africa, the Eastern Mediterranean, and large parts of Asia and Latin America, reinforcing the need for inclusive neuroimaging research.

### Supplemental analysis

Analysis of the Brain Charts Consortium dataset (N = 123,984 scans) revealed that 95.4% originated from HICs, with 3.5% (N = 4,367) from UMICs and 1.1% (N = 1,394) from LMICs. Regional distribution showed predominant representation from EUR (48.4%, N = 59,985) and AMR (44.8%, N = 55,598), together accounting for over 93% of all data. Tamon et al. fMRI meta-analysis demonstrated an even more pronounced level of inequities, with 98.3% of 5,785 cases from HICs and 1.7% (N = 96) from UMICs, with no representation from LMICs or LICs. Regional distribution was similarly restricted: EUR (53.2%, N = 3,078) and AMR (36.6%, N = 2,119) accounted for nearly 90% of cases, with WPR contributing 10.2% (N = 588). Both supplementary datasets contained no data from AFR, EMR, or SEAR regions. These patterns mirror our ENIGMA findings, demonstrating consistent overrepresentation of HICs and systematic underrepresentation of low-resource regions across major neuroimaging initiatives.

## Discussion

Our systematic quantitative synthesis revealed substantial gaps in the global distribution of structural MRI data for mental health and neurological conditions. The overwhelming majority of data (90.5%) originated from HICs, with limited contributions from UMICs (7.9%) and nearly no representation from LMICs (1.6%) or LICs (0%). Geographic representation was similarly limited, with nearly 85% of cases drawn from North America and Europe. At the same time, regions such as Africa, the Eastern Mediterranean, and much of Asia and Latin America were markedly underrepresented. Although some conditions, such as OCD and schizophrenia, demonstrated broader regional spread, most conditions were dominated by data from a narrow set of high-income countries. A supplemental analysis with another consortium and a meta-analysis suggests these results are not specific to ENIGMA and reflect a widespread limitation of the field.

These findings align with broader global data imbalances observed across scientific disciplines, including genetics and epidemiology. For example, in genome-wide association studies (GWAS), approximately 78% of participants are of European descent, while less than 4% represent African, Hispanic, or other ancestries^32^. Structural MRI research shows a similar concentration in Western populations. This lack of diversity limits the clinical relevance and transferability of findings to underrepresented populations and may reinforce existing health inequities. It reflects broader structural imbalances in where research is conducted and by whom, which restricts the scope of scientific conclusions and any associated evidence-informed policy decisions. As Tanaka et al. (2024) show, global collaboration has been increasing to expand structural MRI research across psychiatric and neurological conditions. Their work provides a valuable overview of international MRI initiatives but does not systematically assess distributions by income level, region, or diagnostic category. In contrast, our study provides condition-specific metrics that quantify and map MRI data availability by World Bank income level and WHO region.

These inequities are also present in other areas of medicine. Cardiovascular research, for example, mirrors these inequities. A global analysis of cardiology trials revealed that only 14% of participants were from LMICs and none from LICs, questioning the generalizability of results to highly affected populations^33^. Global cardiovascular risk and mortality estimates often rely on high-income country data, neglecting unique risk factors in low-resource settings^34^. Similar patterns are evident in oncology. Over the last two decades, more than 90% of leukemia clinical trials have been in HICs, with only 4.8% exclusively in LMICs^35^. Furthermore, LMIC trials were significantly less likely to be randomized (43.3% vs 16.3%) and to report overall survival (28.4% vs 47.5%)^35^.

Although our findings use ENIGMA as the primary source of globally harmonized structural MRI data and therefore do not capture the full range of pediatric neuroimaging studies on the mental disorders in focus, they should not be interpreted as a critique of the ENIGMA Consortium itself. On the contrary, ENIGMA represents one of the most inclusive and globally distributed neuroimaging collaborations to date, spanning 47+ countries and involving working group leadership from across Asia, Africa, Latin America, and the Middle East. Many working groups, such as ENIGMA-HIV, ENIGMA-Stroke, ENIGMA-OCD, and ENIGMA-Anxiety, have been co-led or initiated by LMIC-based investigators^11^. Recent initiatives such as ENIGMA-Pakistan^36^, ENIGMA-India^37^, and ENIGMA-Africa further demonstrate how sustained investment through partnerships with funders like the National Institutes of Health and the Fogarty International Center can support data collection in LMICs. In addition, programs like ENIGMA-U extend this impact by offering mentorship and education to neuroscience trainees in over 70 countries, addressing upstream inequities in access to training and career development^38^. Our supplemental analysis further illustrates the need for these efforts. By analyzing data from the Brain Charts Consortium and a large-scale fMRI meta-analysis, we found similarly stark overrepresentation of HICs and persistent underrepresentation of LMICs and entire WHO regions. These consistent patterns across studies reinforce that the disparities identified in ENIGMA reflect systematic, field-wide limitations in global neuroimaging research rather than consortium-specific issues.

### Ethical and developmental considerations

The underrepresentation of LMICs in neuroimaging research has profound implications for children and adolescent mental health globally. Brain development is highly sensitive to environmental factors that vary dramatically across global contexts—including nutritional status, infectious disease burden, educational opportunities, and psychosocial stressors.

Research conducted primarily in HICs may not adequately capture how these environmental factors shape typical and atypical brain development in the majority of the world’s children and adolescents. Furthermore, the absence of neuroimaging data from populations experiencing the highest burden of mental health conditions represents a critical gap in global mental health science. LMICs account for approximately 85% of the world’s youth population, yet current neuroimaging findings may have limited applicability to these populations. This disparity risks perpetuating a cycle where clinical guidelines and interventions developed from HIC data are inappropriately generalized to global contexts The geographic concentration of neuroimaging data raises important ethical questions about scientific responsibility and global health equity^39^. When research findings inform clinical guidelines and treatment recommendations that are applied worldwide, the lack of representation from diverse global populations constitutes more than a methodological limitation—it represents a potential source of health inequity. The scientific community needs to ensure that research evidence reflects the populations to whom findings will be applied.

### Addressing Equity

A critical next step is to systematically identify and address the drivers of underrepresentation in global MRI research. Key structural barriers include the absence of local MRI infrastructure, particularly in settings where even basic clinical neuroimaging is unavailable, raising questions about the feasibility and ethical grounding of advocating for research use. Institutional and political factors are also central, such as the availability of stable governmental or non-governmental partners to support infrastructure investments and the vulnerability of local research programs to shifts in political priorities and funding landscapes. The role of major MRI system manufacturers warrants examination, including their potential responsibilities in promoting equitable global access to neuroimaging technologies. Beyond acquisition, disparities in data sharing infrastructure—ranging from inequities in internet access (e.g., unregulated vs. restricted networks) to the prohibitive costs of cloud services offered by large technology providers—pose substantial challenges. Finally, the harmonization of international data governance frameworks, including divergent data protection regulations, such as Brazil’s Lei Geral de Proteção de Dados (LGPD), the European Union’s General Data Protection Regulation (GDPR), and the United States Health Insurance Portability and Accountability Act (HIPAA)^40^, further complicates cross-border research collaborations.

To build on these efforts and address broader inequities in the field, several strategies should be prioritized. One key strategy involves increased investment in localizing global conferences and capacity-building workshops. As demonstrated by Agustin Ibáñez^41^ and others, hosting scientific meetings in the Global South can help rebalance longstanding Eurocentric structures in science. These gatherings reduce participation barriers, foster trust, build sustainable research networks, and can help to catalyze locally relevant research agendas that are often overlooked in more centralized settings^41^. Efforts like the Genetics of Latin American Diversity (GLAD) Project^42^ demonstrate the critical value of expanding representation to identify population-specific genetic patterns^43^. Additionally, the ReDLat consortium (The multi-partner consortium to expand dementia research in Latin America) provides rich multimodal data, including genetics, neuroimaging, exposome, and plasma biomarkers^4,44,45^.

Scaling these efforts requires consistent and sustained support from global funders. Institutions such as the Wellcome Trust, Global Brain Health Institute, and NIH Fogarty Center have begun to prioritize research equity, yet current investments remain modest relative to global need. The field must advocate for multi-year grants focused on LMIC-led research infrastructure, collaborative networks, and equitable governance models. These actions are not only critical for ethical inclusion, but also necessary to improve the scientific validity and global relevance of neuroimaging research.

### Limitations

There are significant limitations in our study. First, due to the availability of study-level data, the limited number of included conditions restricts the generalizability of our findings across all mental and neurological disorders. Our analysis focused on a subset of nine psychiatric working groups and does not reflect the full scope of ENIGMA’s 50+ active working groups. Including additional groups, particularly in neurology and global health domains, would likely reveal broader geographic participation. Second, we excluded conditions when site-specific data were not reported or when participant counts from multi-country cohorts could not be disaggregated by country, preventing attribution to individual locations. This methodological constraint may have skewed country and regional representation for common mental disorders, such as major depressive disorder and bipolar disorder. Third, limited reporting of age and sex within ENIGMA publications hindered our ability to examine disparities across demographic subgroups. Fourth, although the ENIGMA consortium is globally extensive, it is not the only global neuroimaging dataset, and the conclusions drawn here may not generalize to other datasets. Finally, the relatively small number of studies meeting inclusion criteria limited the statistical comparisons across regions and income levels.

### Conclusion

This systematic quantitative synthesis reveals profound geographic and economic imbalances in the availability of structural MRI data across global mental health research. Despite the international reach and inclusive participatory ethos of the ENIGMA Consortium, our findings show that structural MRI data are overwhelmingly concentrated in high-income countries, with entire regions, including much of Africa and the Eastern Mediterranean, largely absent. These gaps limit the global relevance of neuroimaging science and risk reinforcing disparities in the understanding, preventing, diagnosing, and treating psychiatric and neurological conditions. To promote more equitable research, targeted investment is needed to strengthen neuroimaging infrastructure, support data sharing, and build neuroimaging capacity in low- and middle-income countries. Addressing these structural gaps is critical to ensuring that neuroimaging research reflects and serves the global population.

## Declarations

Human Ethics and Consent to Participate Declarations: Not applicable. Funding: Not applicable. The authors received no specific funding for this work.

## Data Availability

All data analyzed in this study are available online from the original published publicly available sources cited in the manuscript.

## Supplemental Material

**Figure S1.**
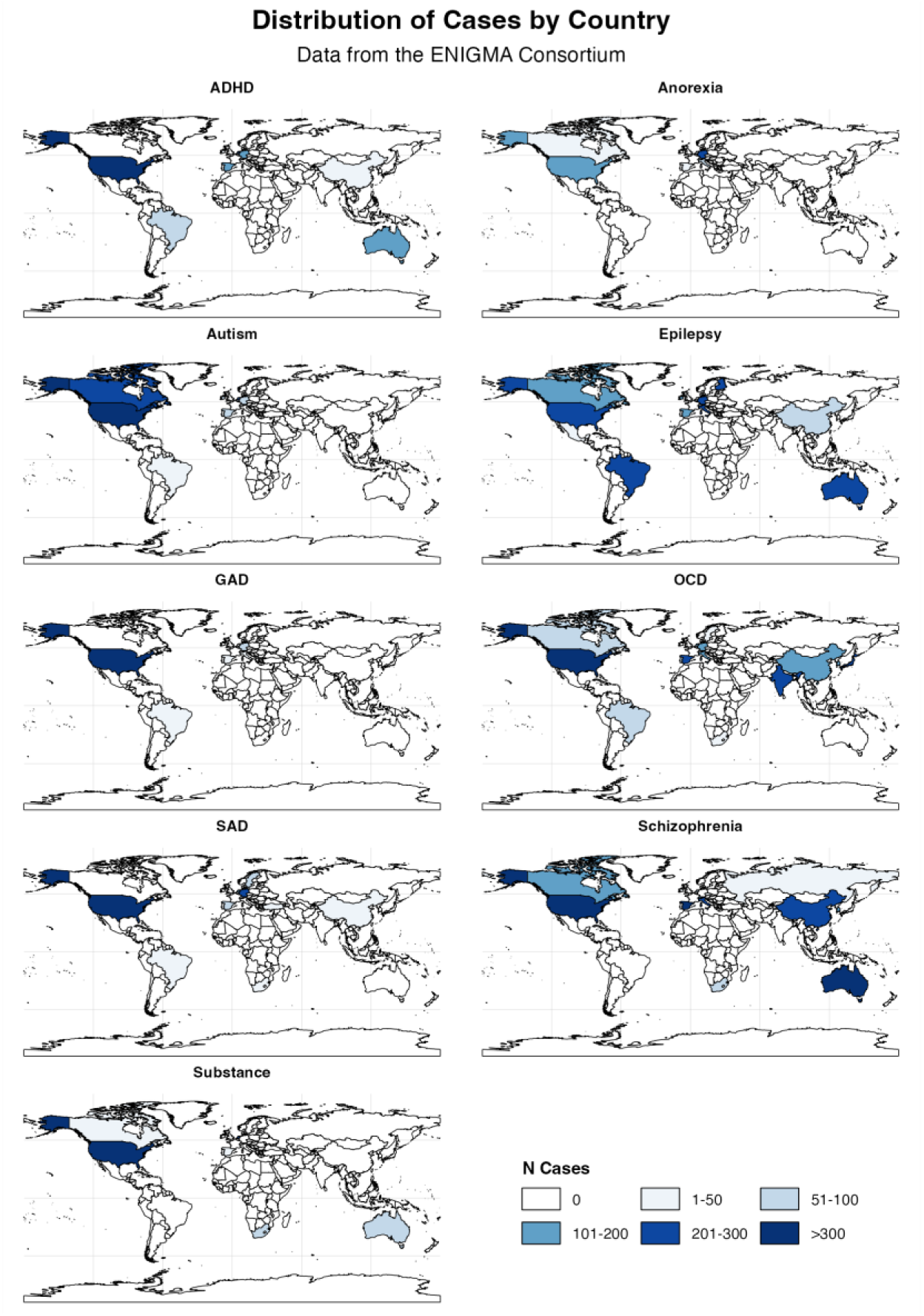
Distribution of Cases by Country. The figure shows the geographic distribution of cases included from the ENIGMA Consortium database across 9 conditions: ADHD, Anorexia, Autism, Epilepsy, Generalized Anxiety Disorder, Obsessive Compulsive Disorder, Social Anxiety Disorder, Schizophrenia, and Substance Use Disorder. Color intensity represents the number of cases reported per country.

**Figure S2.**
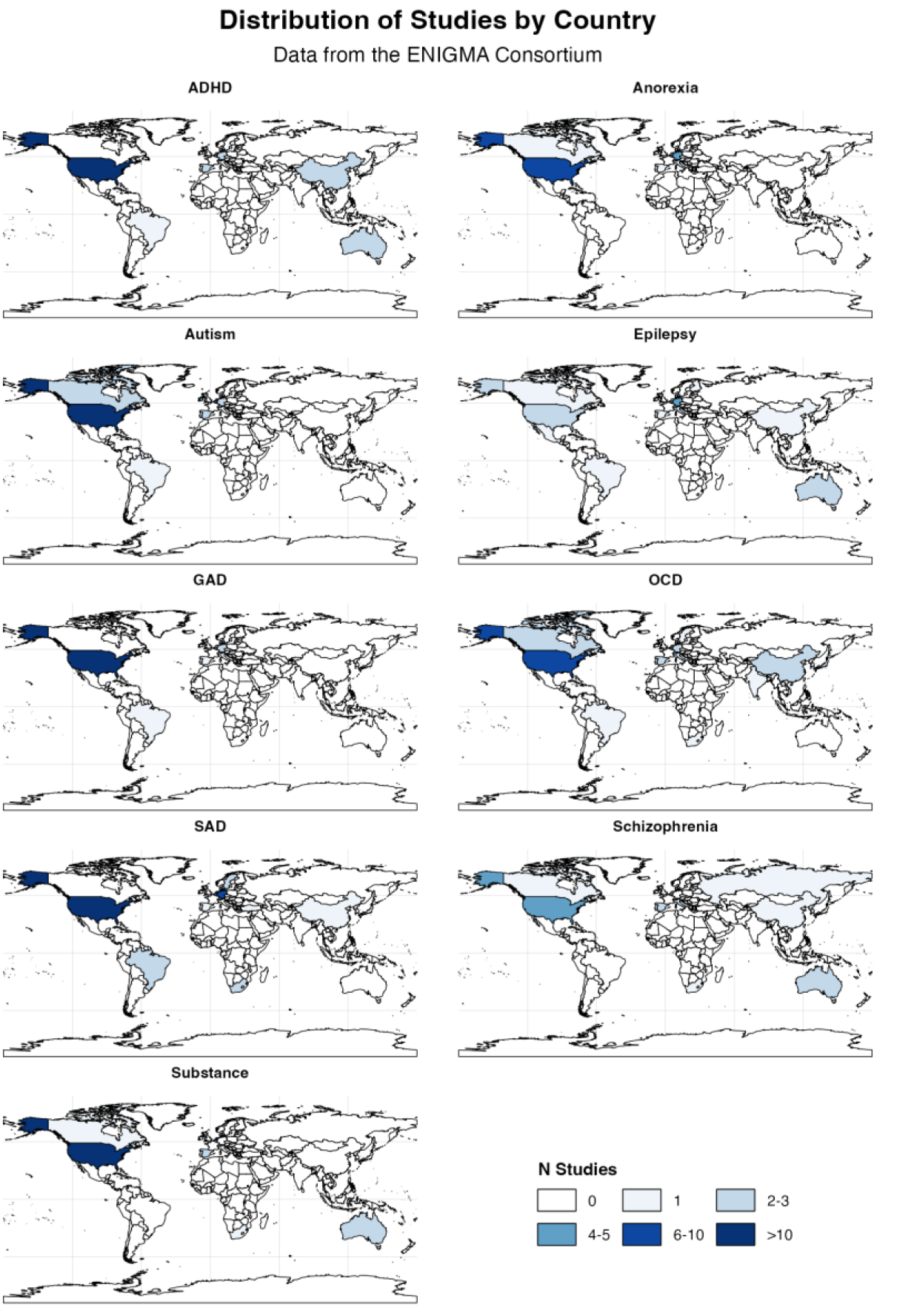
Distribution of Studies by Country. The figure shows the geographic distribution of studies included from the ENIGMA Consortium database across 9 conditions: ADHD, Anorexia, Autism, Epilepsy, Generalized Anxiety Disorder, Obsessive Compulsive Disorder, Social Anxiety Disorder, Schizophrenia, and substance use disorder. Color intensity represents the number of studies reported per country.

**Figure S3.**
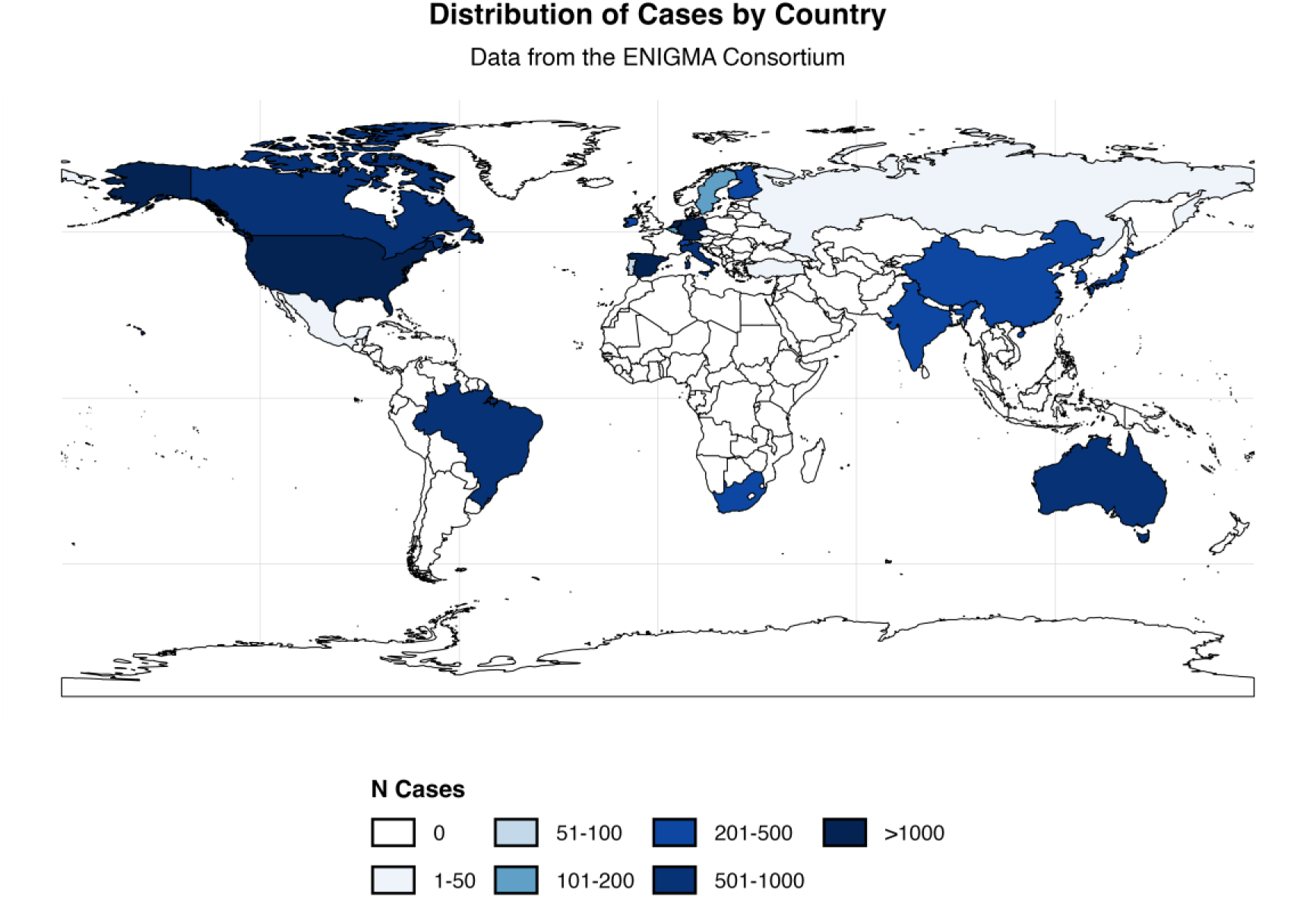
Distribution of Studies by Country. This figure shows the total number of structural MRI cases contributed by each country across all nine ENIGMA conditions. Countries are grouped into distinct categories based on case count: 0, 1–50, 51–100, 101–200, 201–500, 501-1000, and >1000. Darker shades indicate higher case contributions.

**Table S1.** Distribution of case by country, region, and economic status (number of cases, % of all cases).

|  | LIC | LMIC | UMIC | HIC |
| --- | --- | --- | --- | --- |
| **AFR** |  |  | South Africa (207, 1.3%) |  |
| **AMR** |  |  | Brazil (507, 3.2%)<br>Mexico (36, 0.2%) | Canada (649, 4.0%)<br>United States (5996, 37.3%) |
| **EMR** |  |  |  |  |
| **EUR** |  |  | Turkey (30, 0.2%) | Belgium (163, 1.0%)<br>Switzerland (365, 2.3%)<br>Czech Republic (34, 0.2%)<br>Germany (1166, 7.2%)<br>Spain (1247, 7.8%)<br>Finland (240, 1.5%)<br>Ireland (292, 1.8%)<br>Israel (24, 0.1%)<br>Italy (708, 4.4%)<br>Netherlands (1365, 8.5%)<br>Norway (69, 0.4%)<br>Portugal (59, 0.4%)<br>Russian Federation (40, 0.2%)<br>Sweden (116, 0.7%)<br>United Kingdom (514, 3.2%) |
| **SEAR** |  | India (265, 1.6%) |  | South Korea (302, 1.9%) |
| **WPR** |  |  | China (490, 3.0%) | Australia (787, 4.9%)<br>Japan (264, 1.6%)<br>Singapore (151, 0.9%) |

**Table S2.** List of countries and regions according to WHO classification.

| **Region** (number of countries and territories) | **Countries and territories** |
| --- | --- |
| **African Region** (n=47) | Algeria, Angola, Benin, Botswana, Burkina Faso, Burundi, Cabo Verde, Cameroon, Central African Republic, Chad, Comoros, Congo, Côte d'Ivoire, Democratic Republic of the Congo, Equatorial Guinea, Eritrea, Eswatini, Ethiopia, Gabon, Gambia, Ghana, Guinea, Guinea-Bissau, Kenya, Lesotho, Liberia, Madagascar, Malawi, Mali, Mauritania, Mauritius, Mozambique, Namibia, Niger, Nigeria, Rwanda, Sao Tome and Principe, Senegal, Seychelles, Sierra Leone, South Africa, South Sudan, Togo, Uganda, United Republic of Tanzania, Zambia, Zimbabwe. |
| **Region of the Americas** (n=35) | Antigua and Barbuda, Argentina, Bahamas, Barbados, Belize, Bolivia (Plurinational State of), Brazil, Canada, Chile, Colombia, Costa Rica, Cuba, Dominica, Dominican Republic, Ecuador, El Salvador, Grenada, Guatemala, Guyana, Haiti, Honduras, Jamaica, Mexico, Nicaragua, Panama, Paraguay, Peru, Puerto Rico (*Associate WHO Member State), Saint Kitts and Nevis, Saint Lucia, Saint Vincent and the Grenadines, Suriname, Trinidad and Tobago, United States of America, Uruguay, Venezuela (Bolivarian Republic of). |
| **Eastern Mediterranean Region** (n=22) | Afghanistan, Bahrain, Djibouti, Egypt, Iran (Islamic Republic of), Iraq, Jordan, Kuwait, Lebanon, Libya, Morocco, Oman, Pakistan, Qatar, Saudi Arabia, Somalia, Sudan, Syrian Arab Republic, Tunisia, United Arab Emirates, West Bank and Gaza Strip (*Non-Member area), Yemen. |
| **European Region** (n=53) | Albania, Andorra, Armenia, Austria, Azerbaijan, Belarus, Belgium, Bosnia and Herzegovina, Bulgaria, Croatia, Cyprus, Czechia, Denmark, Estonia, Finland, France, Georgia, Germany, Greece, Hungary, Iceland, Ireland, Israel, Italy, Kazakhstan, Kyrgyzstan, Latvia, Lithuania, Luxembourg, Malta, Monaco, Montenegro, Netherlands, North Macedonia, Norway, Poland, Portugal, Republic of Moldova, Romania, Russian Federation, San Marino, Serbia, Slovakia, Slovenia, Spain, Sweden, Switzerland, Tajikistan, Türkiye, Turkmenistan, Ukraine, United Kingdom of Great Britain and Northern Ireland, Uzbekistan. |
| **South-East Asian Region** (n=11) | Bangladesh, Bhutan, Democratic People's Republic of Korea, India, Indonesia, Maldives, Myanmar, Nepal, Sri Lanka, Thailand, Timor-Leste. |
| **Western Pacific Region** (n=28) | Australia, Brunei Darussalam, Cambodia, China, Cook Islands, Fiji, Japan, Kiribati, Lao People's Democratic Republic, Malaysia, Marshall Islands, Micronesia (Federated States of), Mongolia, Nauru, New Zealand, Niue, Palau, Papua New Guinea, Philippines, Republic of Korea, Samoa, Singapore, Solomon Islands, Tokelau (*Associate WHO Member State), Tonga, Tuvalu, Vanuatu, Viet Nam. |

**Table S3.**
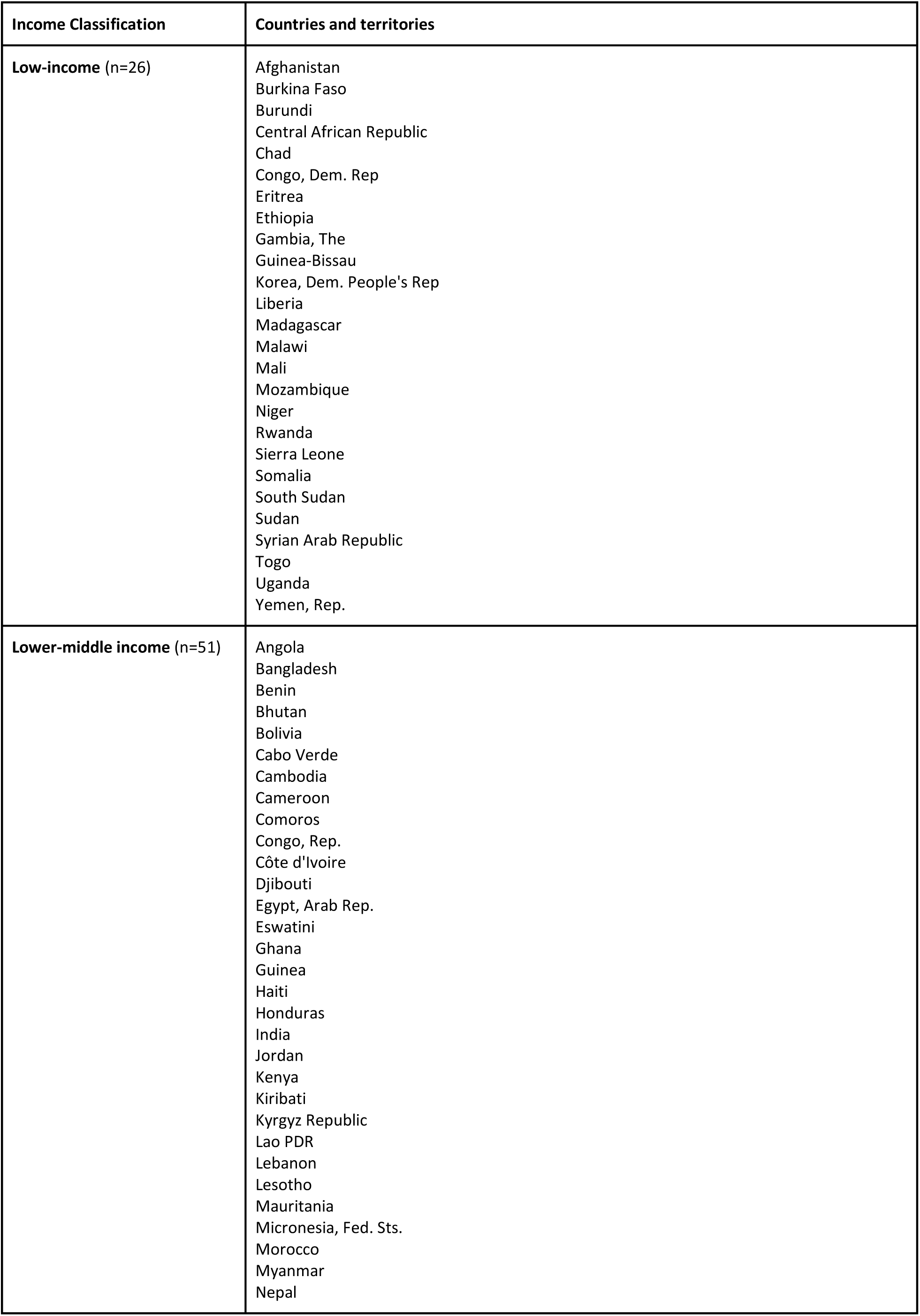
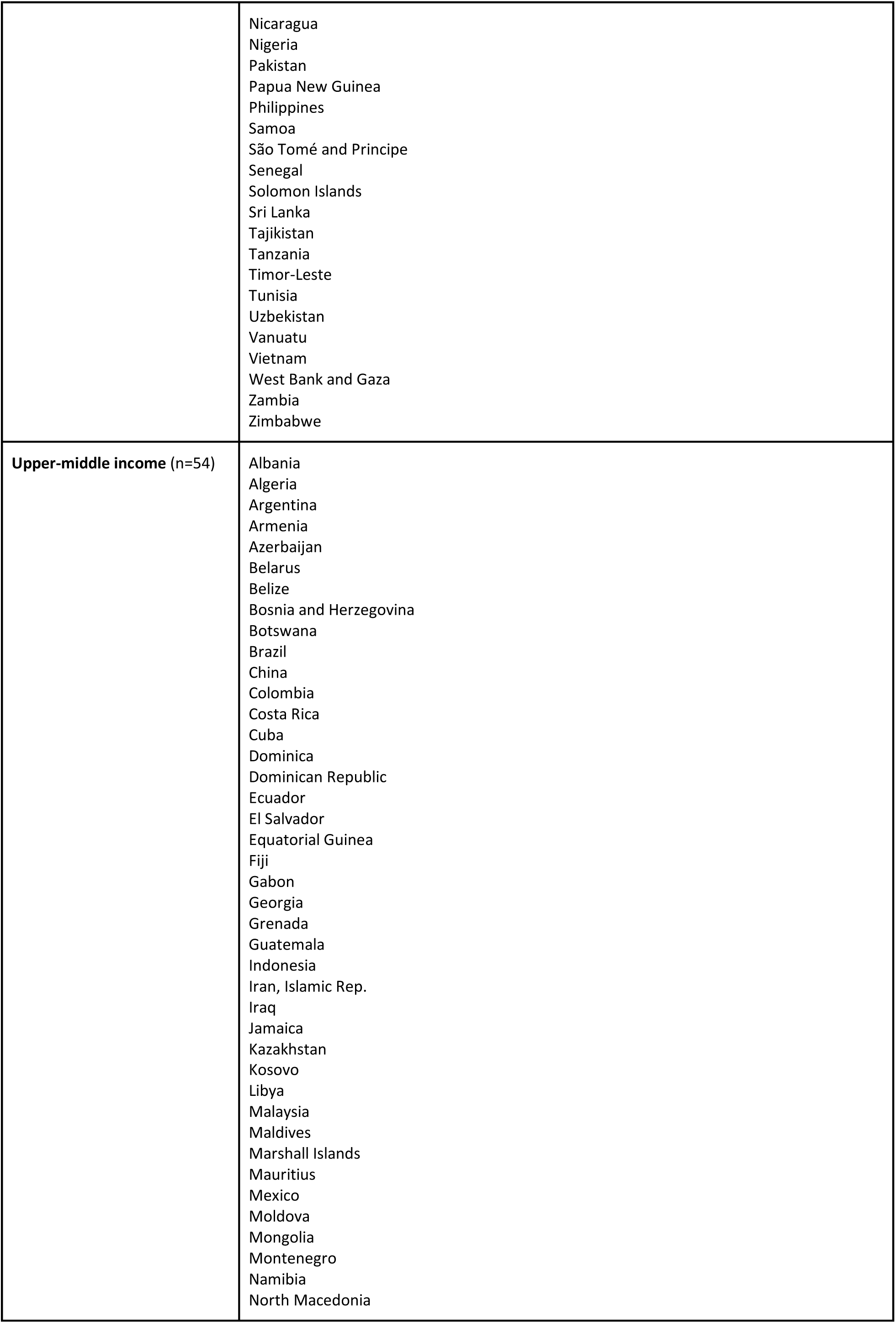
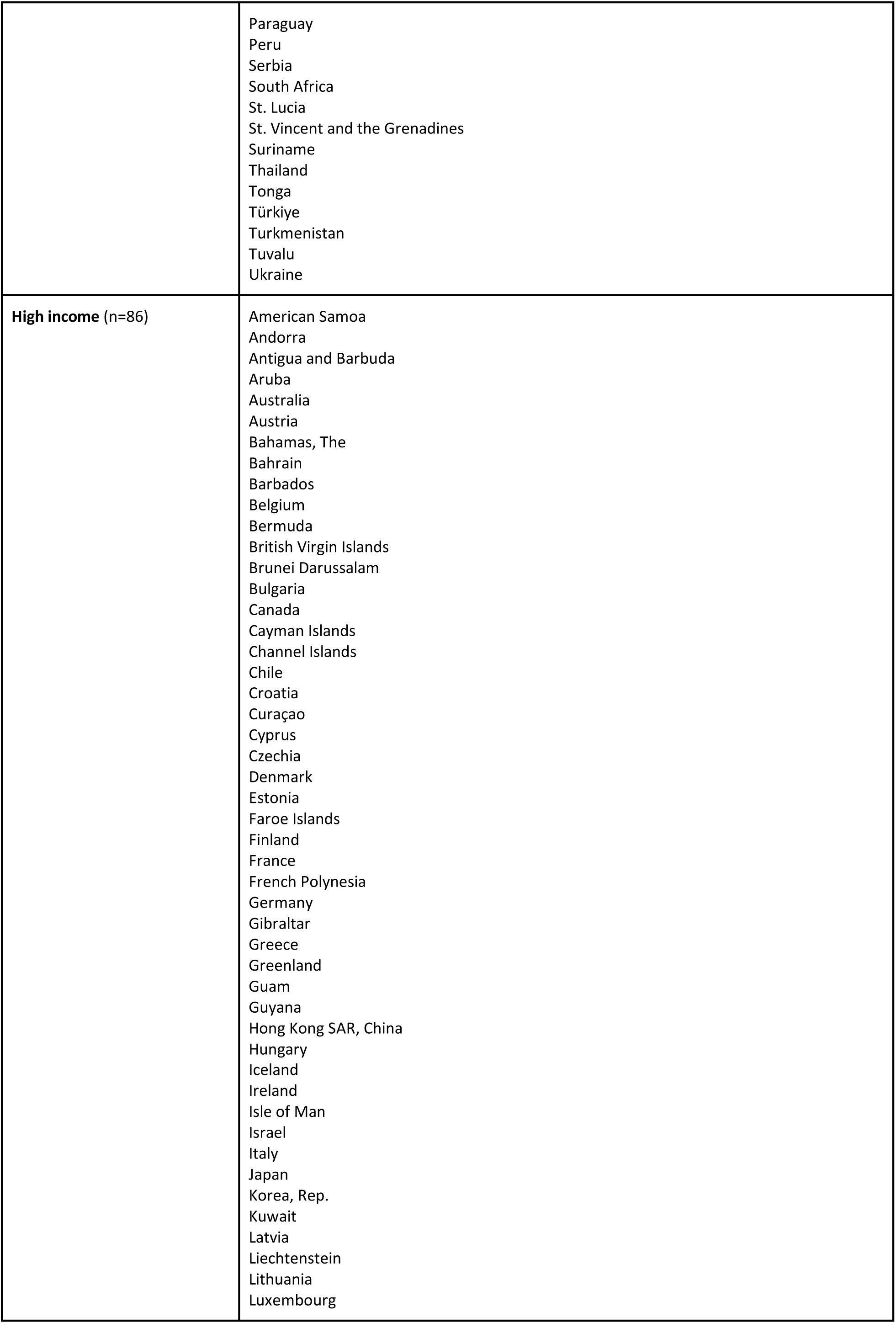
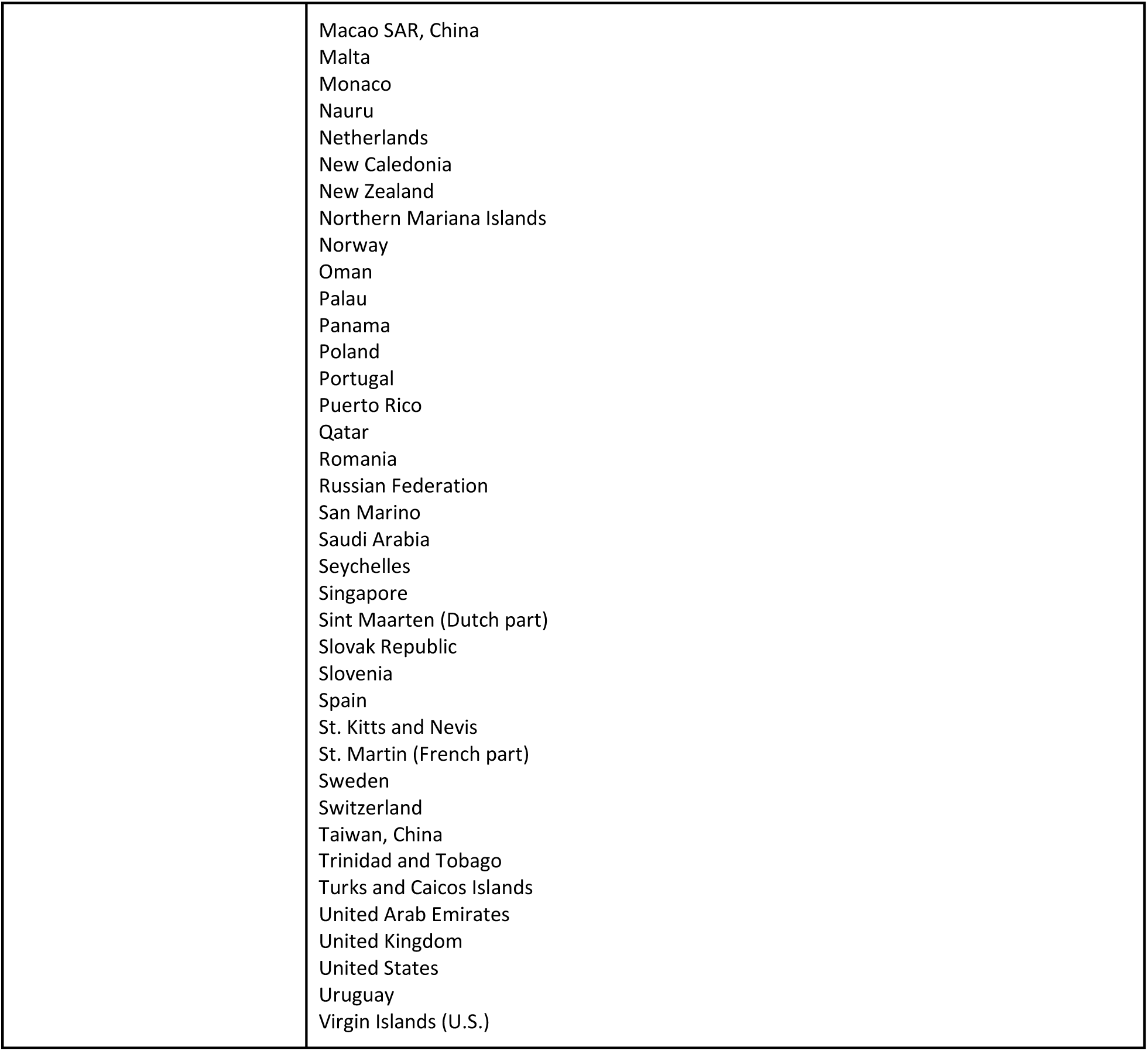
List of countries and regions according to income classification by the World Bank in 2023.

## Notes

### Competing Interest Statement

The authors have declared no competing interest.

### Author Declarations

The study used only openly available, previously published human data from the ENIGMA Consortium and two other publicly accessible datasets: the Brain Charts Consortium and a published task-based fMRI meta-analysis. No new data were collected, and no individual-level or identifiable participant data were accessed. ENIGMA Consortium: https://enigma.ini.usc.edu Brain Charts Consortium (Bethlehem et al., 2022): https://www.nature.com/articles/s41586-022-04554-y fMRI meta-analysis (Tamon et al., 2024): https://pubmed.ncbi.nlm.nih.gov/38685858/

